# Role of IgM and IgA Antibodies in the Neutralization of SARS-CoV-2

**DOI:** 10.1101/2020.08.18.20177303

**Authors:** Jéromine Klingler, Svenja Weiss, Vincenza Itri, Xiaomei Liu, Kasopefoluwa Y. Oguntuyo, Christian Stevens, Satoshi Ikegame, Chuan-Tien Hung, Gospel Enyindah-Asonye, Fatima Amanat, Ian Baine, Suzanne Arinsburg, Juan C. Bandres, Erna Milunka Kojic, Jonathan Stoever, Denise Jurczyszak, Maria Bermudez-Gonzalez, Arthur Nádas, Sean Liu, Benhur Lee, Susan Zolla-Pazner, Catarina E. Hioe

## Abstract

**Background:** SARS-CoV-2 has infected millions of people globally. Virus infection requires the receptor-binding domain (RBD) of the spike protein. Although studies have demonstrated anti-spike and - RBD antibodies to be protective in animal models, and convalescent plasma as a promising therapeutic option, little is known about immunoglobulin (Ig) isotypes capable of blocking infection.

**Methods:** We studied spike- and RBD-specific Ig isotypes in convalescent and acute plasma/sera using a multiplex bead assay. We also determined virus neutralization activities in plasma, sera, and purified Ig fractions using a VSV pseudovirus assay.

**Results:** Spike- and RBD-specific IgM, IgG1, and IgA1 were produced by all or nearly all subjects at variable levels and detected early after infection. All samples displayed neutralizing activity. Regression analyses revealed that IgM and IgG1 contributed most to neutralization, consistent with IgM and IgG fractions’ neutralization potency. IgA also exhibited neutralizing activity, but with lower potency.

**Conclusion:** IgG, IgM and IgA are critical components of convalescent plasma used for COVID-19 treatment.

**Summary of main points:** IgM, IgG1 and IgA1 antibodies against SARS-CoV-2 spike glycoprotein and its receptor-binding domain are present in convalescent COVID-19 plasma. Like IgG, IgM and IgA contribute to virus neutralization, providing the basis for optimal selection of convalescent plasma for COVID-19 treatment.

## Background

Since the first patients with coronavirus disease 2019 (COVID-19), caused by severe acute respiratory syndrome coronavirus 2 (SARS-CoV-2), were identified in Wuhan, China [1], the epidemic has spread worldwide, infecting millions of people. Effective therapeutics and vaccines are urgently needed. Convalescent plasma transfusions have shown promising results in patients with severe COVID-19 [2–4] and clinical trials to evaluate its efficacy for ambulatory and hospitalized patients are underway [5–7]. To this end, information is needed about immunoglobulin (Ig) isotypes in convalescent plasma that have antiviral activities. The data would likewise inform vaccine development [8]. Most vaccines are based on the SARS-CoV-2 spike protein [8,9], which is a membrane-anchored protein present on the virus envelope along with two others (membrane and envelope proteins) and contains the receptor-binding domain (RBD) for binding and entry into cells [10–12]. The vaccines aim to protect by inducing neutralizing antibodies (Abs) that block viral infection.

SARS-CoV-2 spike-, RBD- and nucleocapsid-specific serum and plasma Abs of IgM, IgG, and IgA isotypes are found in most COVID-19 patients [13–18], with neutralizing activities developing within two weeks of infection and declining over time [15,16,19,20]. However, the neutralizing titers vary greatly [15,16,19,20] and correlate with Ab binding levels against RBD, spike, and/or nucleocapsid, and with age, symptom duration, and symptom severity [15,16]. Several RBD-specific monoclonal IgG Abs with neutralizing activity have been generated, and these confer protection in animal models [15,19,21,22]. A monoclonal Ab of IgA isotype recognizing both SARS-CoV-1 and SARS-CoV-2 spike proteins and blocking ACE2 receptor binding was recently described [23]. However, no direct evidence is available regarding the neutralizing capacity of plasma IgM and IgA from COVID-19 patients.

Studies on other respiratory viruses such as influenza show that, in addition to IgG, IgA could also mediate virus neutralization, and their relative contribution depends on the physiologic compartment in which they are found, with IgA contributing to the protection of mostly the upper respiratory tract while IgG was protecting the lower respiratory tract [24,25]. An anti-hemagglutinin monoclonal polymeric IgA has been demonstrated to mediate more potent anti-influenza activities than monoclonal IgG against the same epitope [26]. An IgM monoclonal Ab with neutralizing activity against influenza B has also been described [27]. In addition, respiratory syncytial virus (RSV)-specific mucosal IgA are a better correlate of protection than serum IgG counterparts [28]. In the case of SARS-CoV-1, high titers of IgA in the lungs correlated with reduced pathology in animal models [29]. Whether IgA in the blood and the respiratory tract mucosa offer protection against SARS-CoV-2 remains an open question. Moreover, scant data are available regarding IgM contribution to neutralization and protection against viruses, including SARS-CoV-2. Of note, in terminally ill patients, systemic SARS-CoV-2 infection affects multiple organs [30]. Thus, the capacity of plasma Ig to suppress virus spread is critical for effective therapy against severe COVID-19.

We recently described a multiplex bead Ab-binding assay using the Luminex technology to detect total Ig against spike and RBD [31]. Here we characterized the Ig isotype profiles using the Luminex assay that detects spike- and RBD-specific IgM, IgG1-4, and IgA1-2. Using a pseudovirus assay [32], we also measured plasma or serum neutralization and determined the neutralizing capacity of IgM, IgA, and IgG fractions. The data indicate a high prevalence of spike- and RBD-specific IgM and IgA, similar to that of IgG1, in plasma and sera from COVID-19 patients, and their contributions to virus neutralization. In addition, by testing purified IgG, IgM and IgA fractions from convalescent plasma, this study presents the first direct evidence that plasma IgG, IgM, and IgA all contribute to SARS-CoV-2 neutralization.

## Methods

### Recombinant proteins

SARS-CoV-2 spike and RBD proteins were produced as described [33,34].

### Human samples

All COVID-19-positive and -negative samples tested in this study are tabulated in **Supplementary Table 1**. Twenty-five citrated COVID-19 convalescent plasma samples destined for transfusion to SARS-CoV-2-infected individuals (TF#1-25, collected between March 26^th^ and April 7^th^ 2020) and ten contemporary COVID-19-negative specimens (N#4-13) were obtained from the Division of Transfusion Medicine of the Department of Pathology, Molecular and Cell-Based Medicine (Mount Sinai Hospital System, IRB #20-03574). The convalescent specimens TF#1-25 were from donors pre-screened to have serum IgG reciprocal titer ≥320 in the Mount Sinai Hospital ELISA anti-IgG COVID-19 assay. Four sera from de-identified COVID-19 individuals (P#5-8) were provided by the Clinical Pathology Division of the Department of Pathology, Molecular and Cell-Based Medicine at the Icahn School of Medicine at Mount Sinai. The following samples were obtained from volunteers enrolled in IRB-approved protocols at the Icahn School of Medicine at Mount Sinai (IRB #16-00772, #16-00791, #17-01243) and the James J. Peter Veterans Affairs Medical Center (IRB #BAN-1604): sera from seven participants with documented SARS-CoV-2 infection (P#1 d8, d11, and d15 after symptom onset, P#2 d7 and d10 after symptom onset, and RP#1-5 after convalescence), and pre-pandemic sera from twelve healthy donors (N#1-3, N#14-22). All study participants provided written consent. All samples were heat-inactivated before use.

### Ig fractionation

IgA was isolated first from plasma using peptide M agarose beads (InvivoGen #GEL-PDM). The pass-through plasma was enriched sequentially for IgG using protein G agarose beads (InvivoGen #GEL-AGG) and for IgM using a HiTrap IgM column (G.E. Healthcare #17-5110-01). An additional step was performed using Protein A Plus mini-spin columns to separate IgG from IgM. Protein concentrations were determined with Nanodrop (Thermo Scientific).

### Multiplex bead Ab binding assay

SARS-CoV-2 spike and RBD antigens were coupled to beads and experiments performed as described [31] except for the use of different secondary Abs designated in the figure legends.

### COV2pp production and titration

SARS-CoV-2 pseudoviruses (COV2pp) with wild-type (WT) or D614G-mutated spike proteins were produced as described [32]. Pseudoviruses were titrated on 20,000 Vero-CCL81 cells seeded 24 hours before infection. At 18-22 hours post-infection, the infected cells were washed and Renilla luciferase activity was measured with the Renilla-Glo™ Luciferase Assay System (Promega #E2720) on a Cytation3 (BioTek) instrument.

### COV2pp neutralization

Virus was pre-incubated with diluted samples for 30 minutes. The virus-sample mix was then added to Vero-CCL81 cells seeded 24 hours earlier and spinoculated. Infection was measured after 18-22 hours by luciferase activity.

The percentage of neutralization was calculated as follows: 100-([sample RLU - cell control RLU]/virus control RLU)*100. IC_50_ and IC_90_ titers were calculated as the reciprocal sample dilution or purified Ig fraction concentration achieving 50% and 90% neutralization, respectively.

### Statistical analysis

Two-tailed Mann-Whitney test, Spearman rank-order correlation test, and simple linear regressions were performed as designated in the figure legends using GraphPad Prism 8.

## Results

### Levels of Ig isotypes against the SARS-CoV-2 spike and RBD vary in convalescent individuals

A total of 29 serum (P#5-8) and plasma (TF#1-25) specimens from COVID-19 convalescent individuals was tested. TF#1-25 were collected ∼4-8 weeks after the initial outbreak in North American, and used for transfusion into hospitalized COVID-19 patients [2]. Ten plasma from COVID-negative contemporaneous blood bank donors (N#4-13) were included for comparison. Sera or plasma from 12 uninfected individuals banked prior to the COVID-19 outbreak (N#1-3 and N#14-22) were used to establish background values. The specimens were initially titrated for total Ig against spike and RBD (**Fig. 1**). All 29 COVID-19 positive specimens exhibited titration curves of total Ig Abs against spike, while none of the negative controls displayed reactivity. Similar results were observed with RBD, except that one contemporaneous COVID-19-negative sample had a low level of RBD-specific Ig (N#10). Overall, the background MFI values were higher for RBD than spike. To assess the reproducibility of the assay, the samples were tested in at least two separate experiments run on different days, and a strong correlation was observed between the MFI values from these independent experiments (**Supplementary Fig. 1**). The areas under the curves (AUCs) highly correlated with the MFI values from specimens diluted 1:200 (p <0.0001; **Supplementary Fig. 2**); consequently, all samples were tested for isotyping at this dilution. At the 1:200 dilution we were able to discern a diverse range of Ig isotype levels among individual samples (**Fig. 2**). To evaluate for the presence of spike-specific and RBD-specific total Ig, IgM, IgG1, IgG2, IgG3, IgG4, IgA1 and IgA2, the specificity and strength of the secondary Abs used to detect the different isotypes were first validated with Luminex beads coated with myeloma proteins of known Ig isotypes (IgG1, IgG2, IgG3, IgG4, IgA1, IgA2, and IgM). All eight secondary Abs were able to detect their specific Ig isotypes with MFI values reaching >60,000 (**Supplementary Fig. 3**).

**Fig 1.**
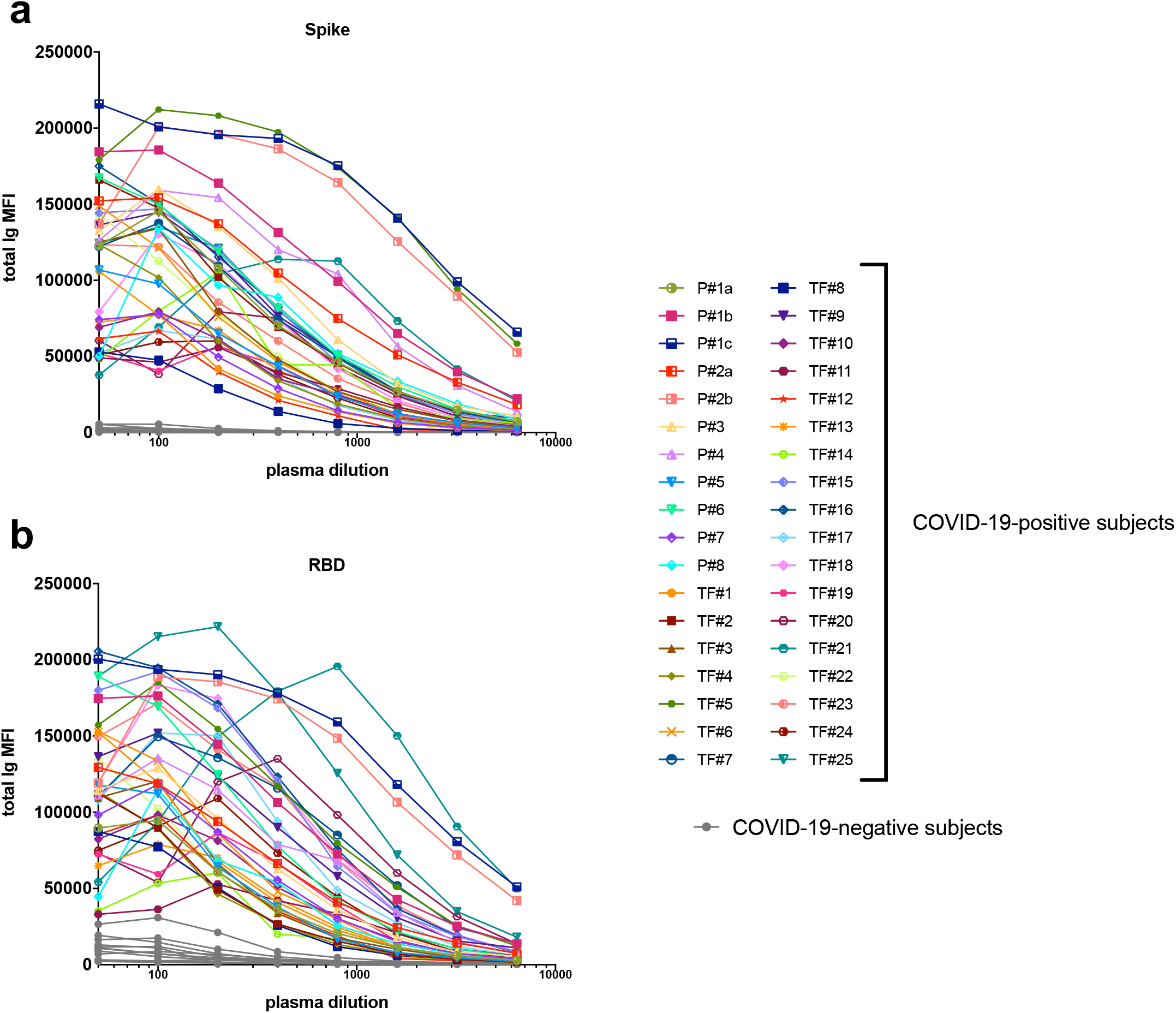
Titration of SARS-CoV-2 spike and RBD total Ig in plasma or serum samples from COVID-19 convalescent individuals. Titration of (a) spike-specific or (b) RBD-specific total Ig from 29 COVID-19 convalescent individuals, two acute COVID-19 patients with longitudinal samples, and 13 COVID-19 uninfected negative individuals. Specimens were diluted at 2-fold dilutions from 1:50 to 1:6,400.

**Fig 2.**
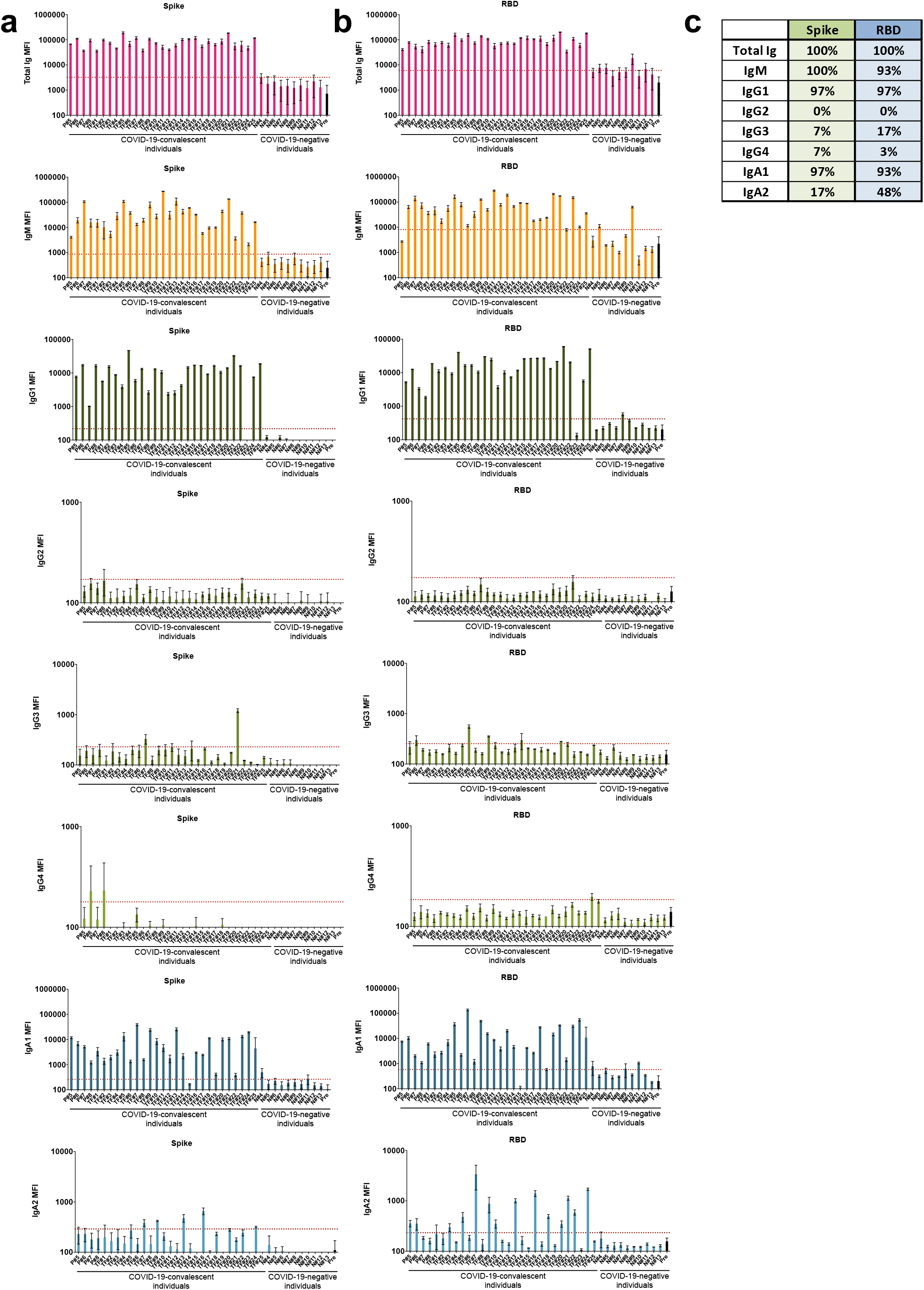
Levels of Ig isotypes against the SARS-CoV-2 spike and RBD vary in plasma or serum samples from COVID-19 convalescent individuals. Total Ig, IgM, IgG1, IgG2, IgG3, IgG4, IgA1 and IgA2 against (a) spike and (b) RBD in specimens from 29 COVID-19 convalescent individuals, 13 COVID-19 uninfected contemporaneous samples, and pre-pandemic controls were detected using the following secondary Abs: rabbit biotinylated-anti-human total Ig (Abcam, catalog #ab97158) at 2 μg/mL, mouse biotinylated-anti-human IgG1 Fc (Invitrogen #MH1515) at 4 μg/mL, mouse biotinylated-anti-human IgG2 Fc (Southern Biotech #9060-08) at 1 μg/mL, mouse biotinylated-anti-human IgG3 Hinge (Southern Biotech #9210-08) at 3 μg/mL, mouse biotinylated-anti-human IgG4 Fc (Southern Biotech #9200-08) at 4 μg/mL, mouse biotinylated-anti-human IgA1 Fc (Southern Biotech #9130-08) at 4 μg/mL, mouse biotinylated-anti-human IgA2 Fc (Southern Biotech #9140-08) at 4 μg/mL or goat biotinylated-anti-human IgM (Southern Biotech #2020-08) at 3 μg/mL. The samples were tested at a dilution of 1:200 and data are shown as mean MFI + standard deviation (SD) of duplicate measurements from at least two independent experiments. The pre-pandemic controls are shown as mean MFI + SD of 12 samples (Pre, black bar). The horizontal red dotted line represents the cut-off value determined as the mean + 3 SD of 12 pre-pandemic samples for each of the isotypes. (c) Percentages of responders above the cut-off for each spike- or RBD-specific Ig isotype.

**Fig 3.**
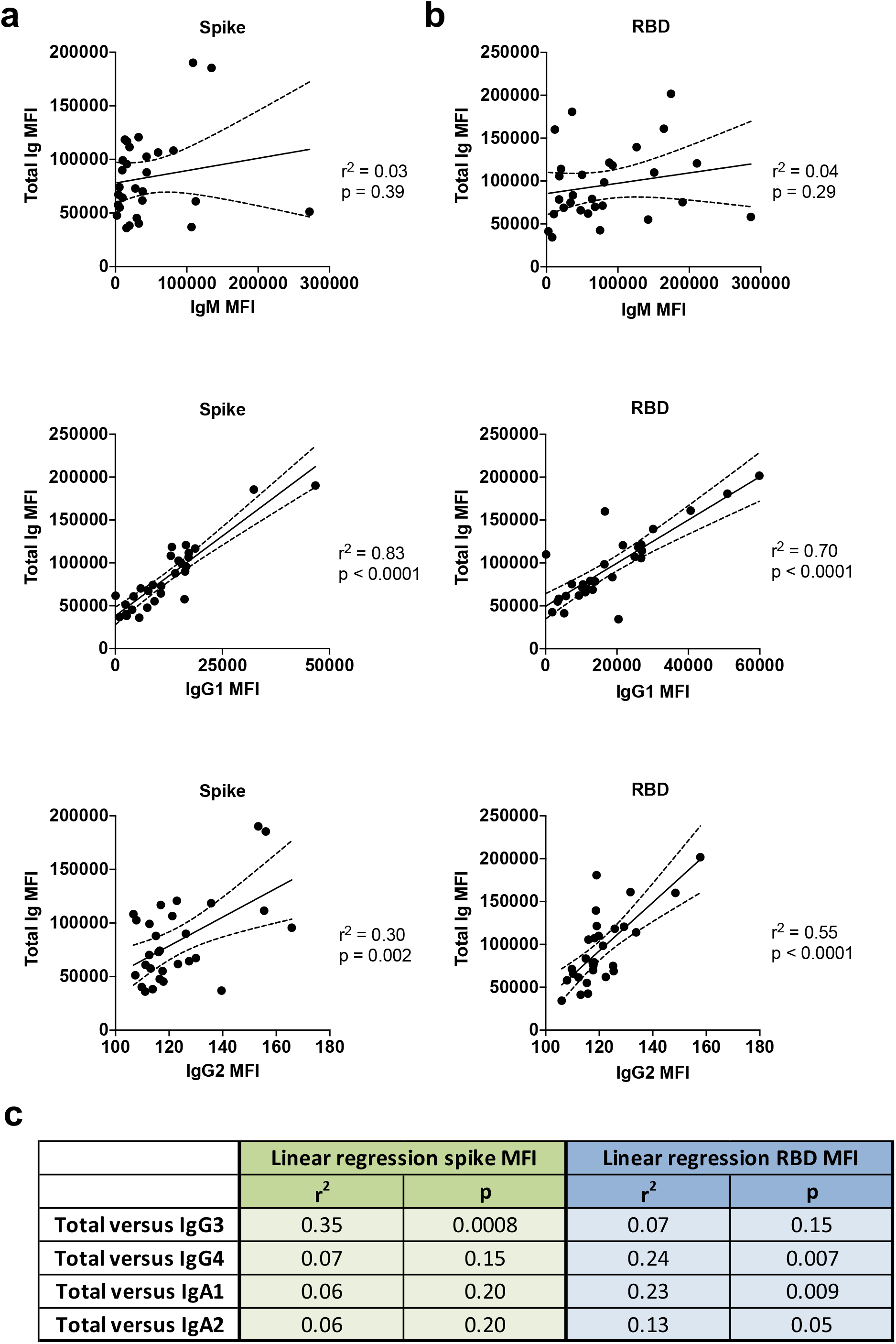
IgG1 is the dominant isotype induced in COVID-19 convalescent individuals. Simple linear regression of (a) spike-specific or (b) RBD-specific total Ig levels versus IgM, IgG1 or IgG2 levels versus (c) spike-specific and RBD-specific IgG3, IgG4, IgA1, and IgA2 levels from the 29 COVID-19-convalescent individuals from Fig. 1. The dash lines represent 95% confidence intervals.

All 29 convalescent individuals had anti-spike and anti-RBD total Ig (**Fig. 2**), but the Ig levels were highly variable, with MFI values ranging from 36,083 to 190,150. In addition, all 29 convalescent individuals also displayed IgM Abs against spike at varying levels, and 93% were positive for anti-RBD IgM when evaluated using cut-off values calculated as mean + 3 standard deviation (SD) of the 12 pre-pandemic samples (**Fig 2b, c**). An IgG1 response was detected against both spike and RBD in 97% of the convalescent subjects, with MFI values that ranged from 1,013 to 59,880. In contrast, IgG2, IgG3, and IgG4 Abs against spike and RBD were detected in only a small fraction of the subjects, and the levels were very low (MFI <1,300) (**Fig. 2**). Surprisingly, almost all individuals produced IgA1 Abs against spike (97%) and RBD (93%), while 17% exhibited IgA2 against spike, and 48% exhibited IgA2 against RBD (**Fig. 2**). Low levels, slightly above cut-off, of spike- and RBD-binding total Ig, IgM, IgG1, and IgA1 were detected sporadically in contemporaneous COVID-19 samples, such as N#8, N#10, and N#11. The responses against spike and RBD were highly correlated for every isotype (**Supplementary Fig. 4**). Overall, these data demonstrate that IgM, IgG1, and IgA1 Abs were induced against spike and RBD in all or almost all COVID-19 convalescent individuals (**Fig. 2**). The levels, however, were highly variable among individuals. No significant difference was observed between female and male individuals (**Supplementary Fig. 5**).

**Fig 4.**
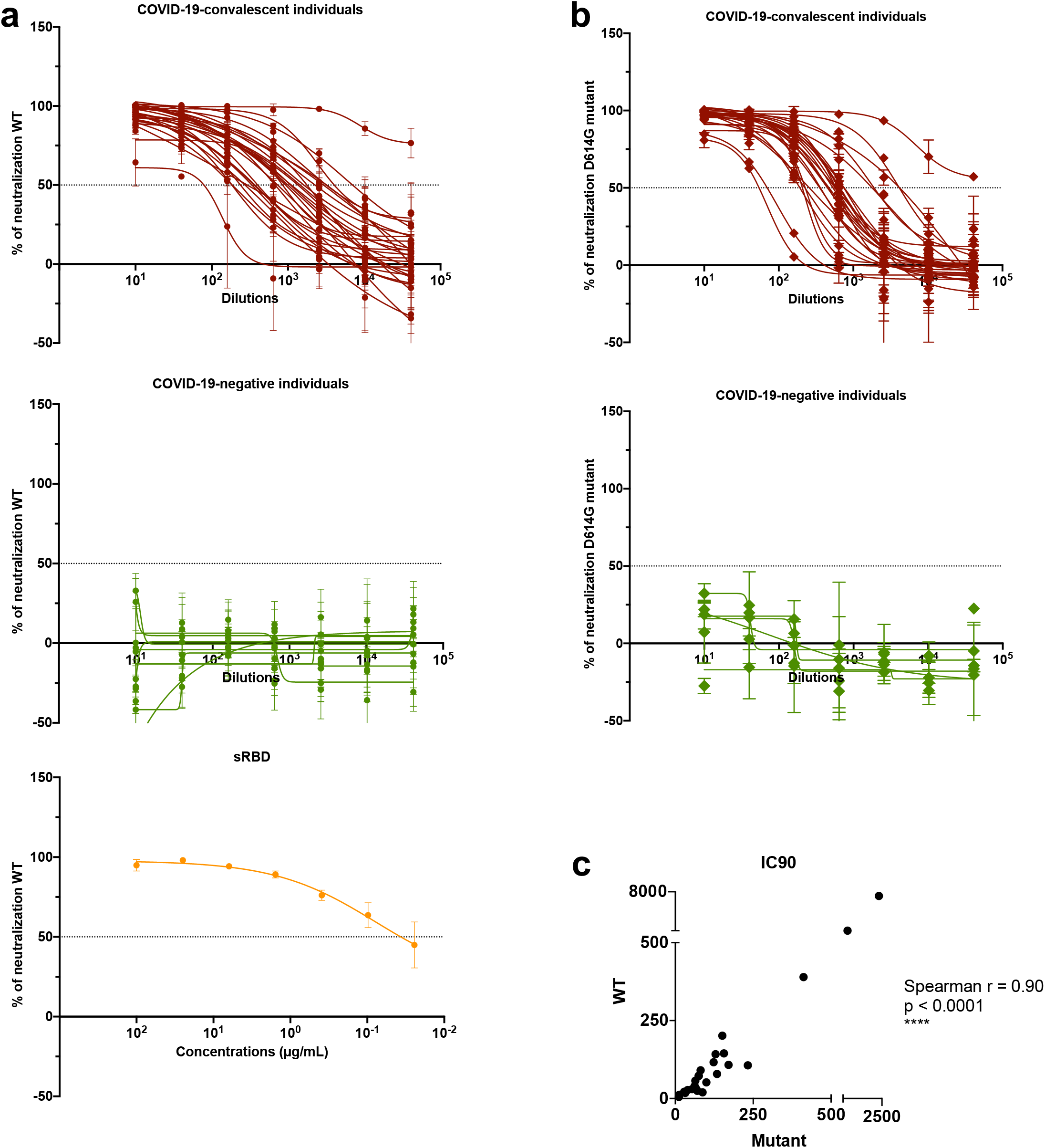
Neutralization activities are detected in all COVID-19 convalescent individuals. Neutralization of COV2pp with (a) WT or (b) D614G mutated spike proteins by samples from 28 COVID-19convalescent individuals and 11 COVID-19 uninfected individuals. The neutralizing activity of recombinant soluble RBD (sRBD) is shown as a positive control. Twenty-four hours before infection, 20,000 Vero-CCL81 cells/well were seeded. Virus (82.5 µL/well) was pre-incubated with serially diluted samples (27.5 µL/well, 4-fold from 1:10 to 1:40,960) for 30 minutes at room temperature. The virus/sample mix was then added to the cells and spinoculated by centrifugation (1250 rpm, 1 hour, room temperature). Six virus-only and six medium-only wells were kept for each plate. After 18 to 22 hours at 37^°^C, infection was measured by luciferase activity. sRBD was tested as a positive control at 4-fold dilutions from 100 to 0.02 µg/mL. The data are shown as mean percentage of neutralization + SD of triplicates. The extrapolated titration curves were generated using a nonlinear regression model in GraphPad Prism (Inhibitor versus response – variable slope [four parameters], least squares regression). The dotted horizontal lines highlight 50% neutralization. (c) Spearman correlation between the IC_90_ titers against COV2pp WT vs. D614G.

**Fig 5.**
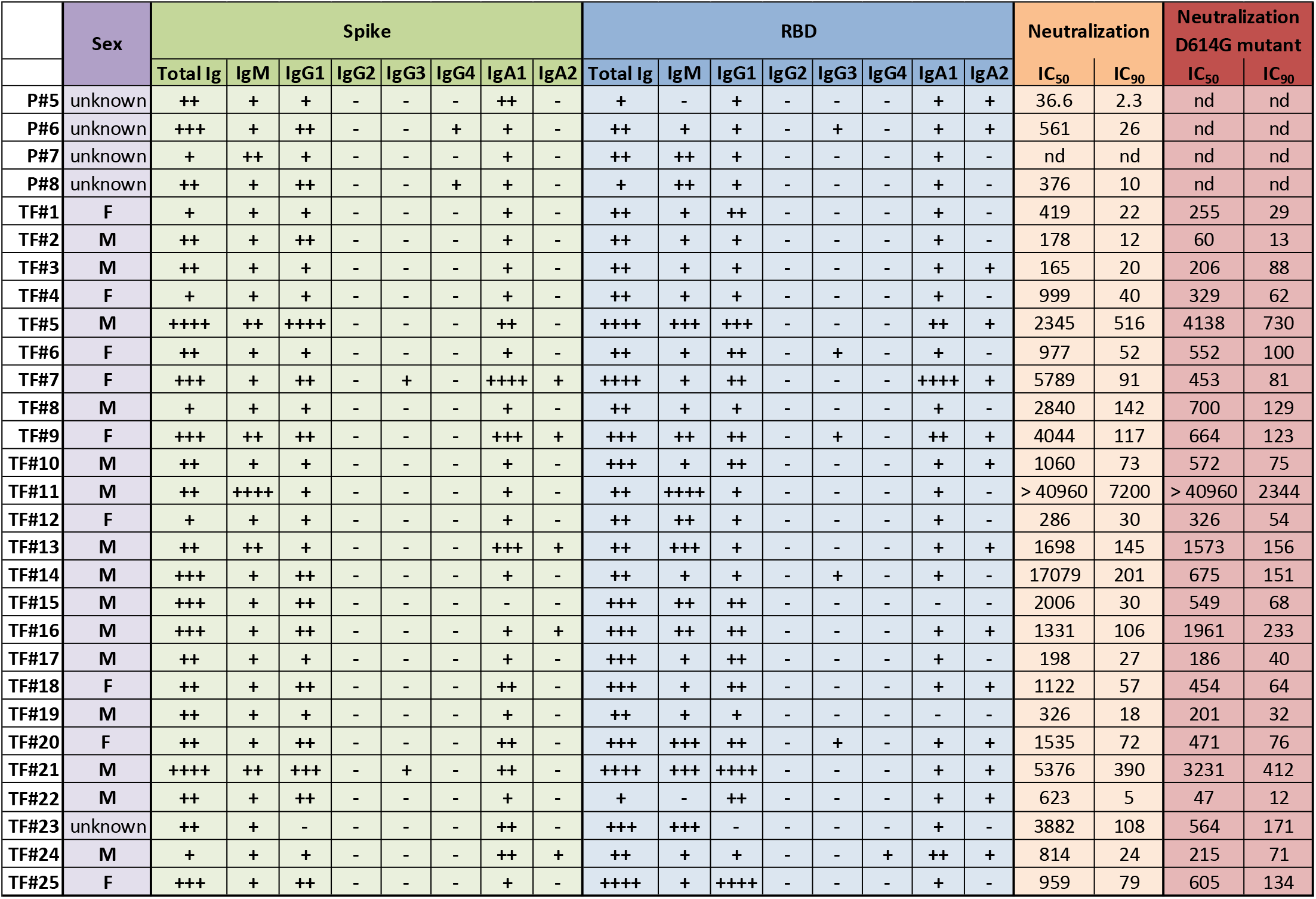
Summary of relative Ig isotype levels and neutralization titers. Table showing sex (purple, F: female, M: male), relative levels of spike-specific (green) and RBD-specific (blue) Ig isotypes (+: bottom quartile, ++: second quartile, +++: third quartile, ++++: top quartile, -: non-responder) and reciprocal IC_50_ and IC_90_ neutralization titers against WT pseudovirus (orange) and D614G pseudovirus (red) of 29 plasma samples from COVID-19 convalescent individuals. nd: not done.

In **Fig. 3**, regression analyses to assess the impact of individual isotypes on the total Ig binding showed that IgG1 had the highest r^2^ values (0.83 and 0.70 for spike- and RBD-binding IgG1, respectively) with p <0.0001, indicating that IgG1 is the major isotype induced by SARS-CoV-2 infection against spike and RBD (**Fig. 3a,b**). IgG2 Abs against RBD had an r^2^ value of 0.55 with p <0.0001, but IgG2 levels were very low. For all other isotypes, including IgM, the r^2^ values were <0.40 (**Fig. 3c**). Thus, despite the presence of many isotypes in sera and plasma, as expected, the major isotype of spike and RBD-specific Abs is IgG1.

Specimens from two patients (P#1 and P#2) were drawn during the acute phase of the infection. Serial specimens from these patients were tested to determine the isotypes of Abs present early in infection. The earliest samples from both patients, drawn at 7 or 8 days after symptom onset were already positive for total Ig, IgG1, IgA1and IgM Abs against spike and RBD (**Supplementary Fig. 6**), and these levels increased over the following three to seven days. On the contrary, IgA2 Ab levels were near or below background on days 7-8 and remained unchanged over the two weeks post-onset. IgG4 Abs also remained low or near background, whereas IgG2 and IgG3 Abs increased slightly to above background after 10-15 days.

**Fig 6.**
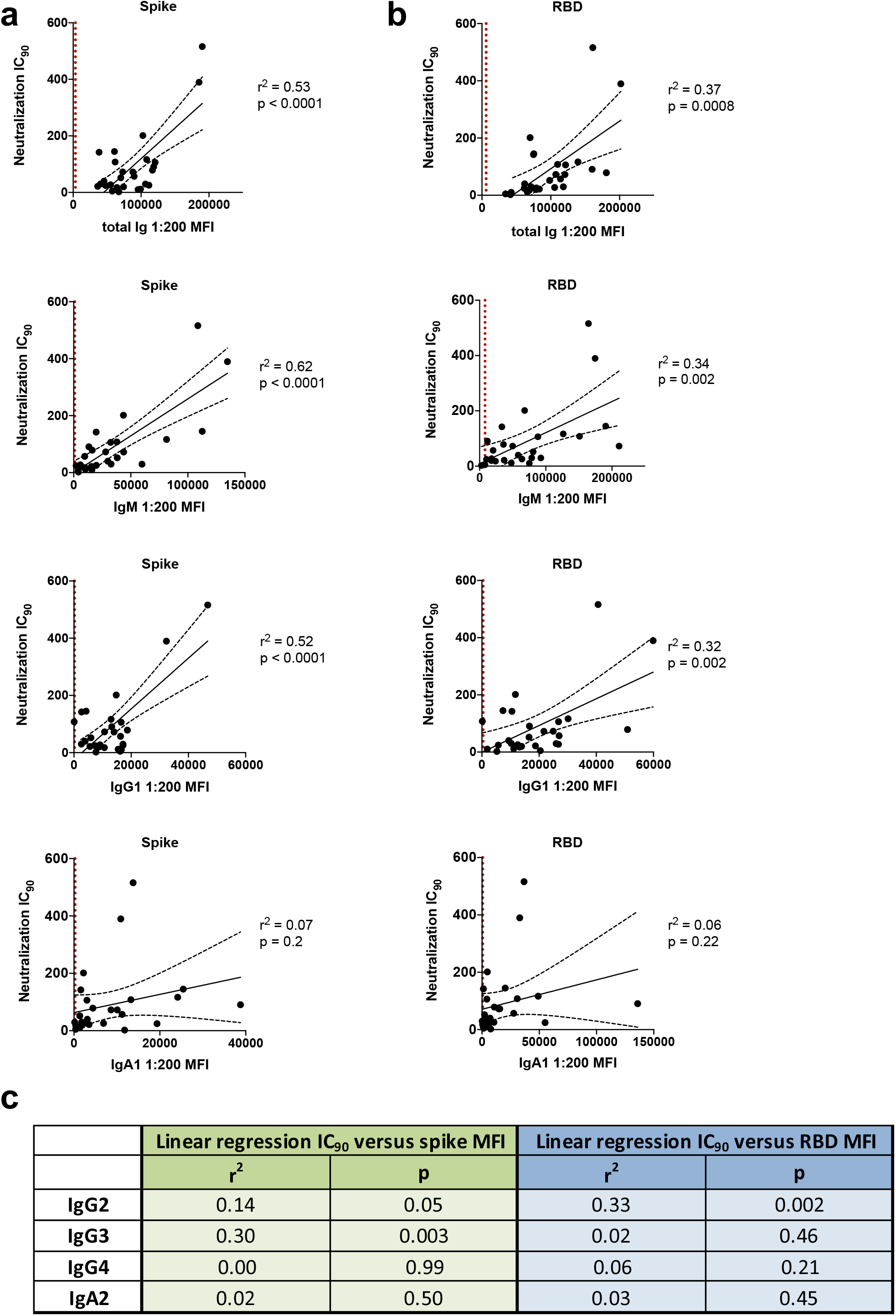
IgM and IgG1 contribute most to SARS-CoV-2 neutralization. Simple linear regression of reciprocal IC_90_ neutralization titers of 27 COVID-19 convalescent individuals versus (a) spike-specific or (b) RBD-specific total Ig, IgM, IgG1 and IgA1 Ab levels. The black dash lines show the 95% confidence intervals. The dotted vertical red line represents the cut-off (mean of 12 pre-pandemic samples + 3 SD) for each isotype from Fig. 1. (c) Statistical results of simple linear regression analyses of reciprocal IC_90_ neutralization titers of 27 COVID-19 convalescent individuals versus spike-specific or RBD-specific Abs levels for IgG2-4 and IgA2.

### Neutralizing activity is detected in specimens from all convalescent COVID-19 individuals

We subsequently tested the ability of samples from convalescent subjects to neutralize a VSVΔG pseudovirus bearing the SARS-CoV-2 spike protein (COV2pp). This pseudovirus assay demonstrated a strong positive correlation with neutralization of the authentic SARS-CoV-2 virus [32]. The titration of neutralizing activity against the WT COV2pp is shown in **Fig. 4a** for specimens from 28 COVID-19 convalescent individuals and 11 uninfected individuals, tested over a range of seven serial four-fold dilutions. A soluble recombinant RBD (sRBD) protein capable of blocking virus infection was tested in parallel as a positive control.

All specimens from COVID-19 convalescent individuals were able to neutralize the virus at levels above 50% (**Fig. 4a**). For 26 of 28 specimens, neutralization reached >90% (**Fig. 4a**). The sample with the lowest titer (reciprocal IC_50_ titer = 37) reached a neutralization plateau of only ∼60%. Of note, one sample (TF#11) demonstrated highly potent neutralization with a reciprocal IC_50_ titer > 40,960, and neutralization was still 75% at the highest dilution tested. None of the samples from uninfected individuals reached 50% neutralization (**Fig. 4a**), while the sRBD positive control demonstrated potent neutralization with an IC_50_ of 0.06 µg/mL (**Fig. 4a**), similar to that recently reported [32].

The samples were also tested for neutralization against a COV2pp bearing the spike with a D614 mutation (D614G mutant), as the D614G variant has become the most prevalent circulating strain in the global pandemic [35]. Similar to the WT COV2pp, all COVID-19-convalescent samples had neutralizing activity reaching >50%, while none of the negative samples did (**Fig. 4b**). The IC_90_ titers against WT and D614 mutant differed on average by only 1.7-fold and correlated strongly with each other (p<0.0001, **Fig. 4c**).

### IgM and IgG1 contribute most to SARS-CoV-2 neutralization

Given our observation that Ab isotype levels and neutralization titers varied tremendously among convalescent COVID-19 individuals (**Figs. 2 and 5**), we investigated the relative contribution of each Ab isotype to the neutralizing activities. Regression analyses were performed on 27 COVID-19 convalescent samples (TF#11 was excluded due to its outlier neutralization titer). As expected, relatively high r^2^ values (0.32–0.62) and significant p values were observed with total Ig, IgM and IgG1; in each case, r^2^ values were higher for spike than for RBD (**Fig. 6a**). The highest r^2^ value was achieved in the analysis of IC_90_ neutralizing titers and IgM binding to spike (r^2^=0.62). For other isotypes, significant p values were sporadically achieved, but r^2^ values were weak (**Fig. 6a,b**).

### Neutralizing activities are mediated by plasma IgM, IgG, and IgA fractions

To assess directly the capacity of different isotypes to mediate neutralization, we evaluated the neutralization activities of IgM, IgG, and IgA fractions purified from the plasma of five COVID-19 convalescent individuals (RP#1-5). The enrichment of IgM, IgG1, and IgA1 Abs reactive with spike and RBD was validated using the isotyping method used above (**Supplementary Fig. 7** and data not shown). These IgM, IgG, and IgA fractions were then evaluated for neutralizing activity along with the original plasma (**Fig. 7**). The RP#1-5 plasma neutralizing reciprocal IC_50_ titers ranged from 35 to 690 (**Fig. 7a,b**). Purified IgM and IgG fractions from RP#1-5 all mediated neutralization reaching more than 50%. Unexpectedly, plasma IgA fractions also displayed neutralizing activity, although not with the same potency as IgM and IgG (**Fig 7c,d**). In contrast, IgM, IgG, and IgA fractions from the negative control (RN#1) showed no neutralization (**Fig. 7c,d**).

**Fig 7.**
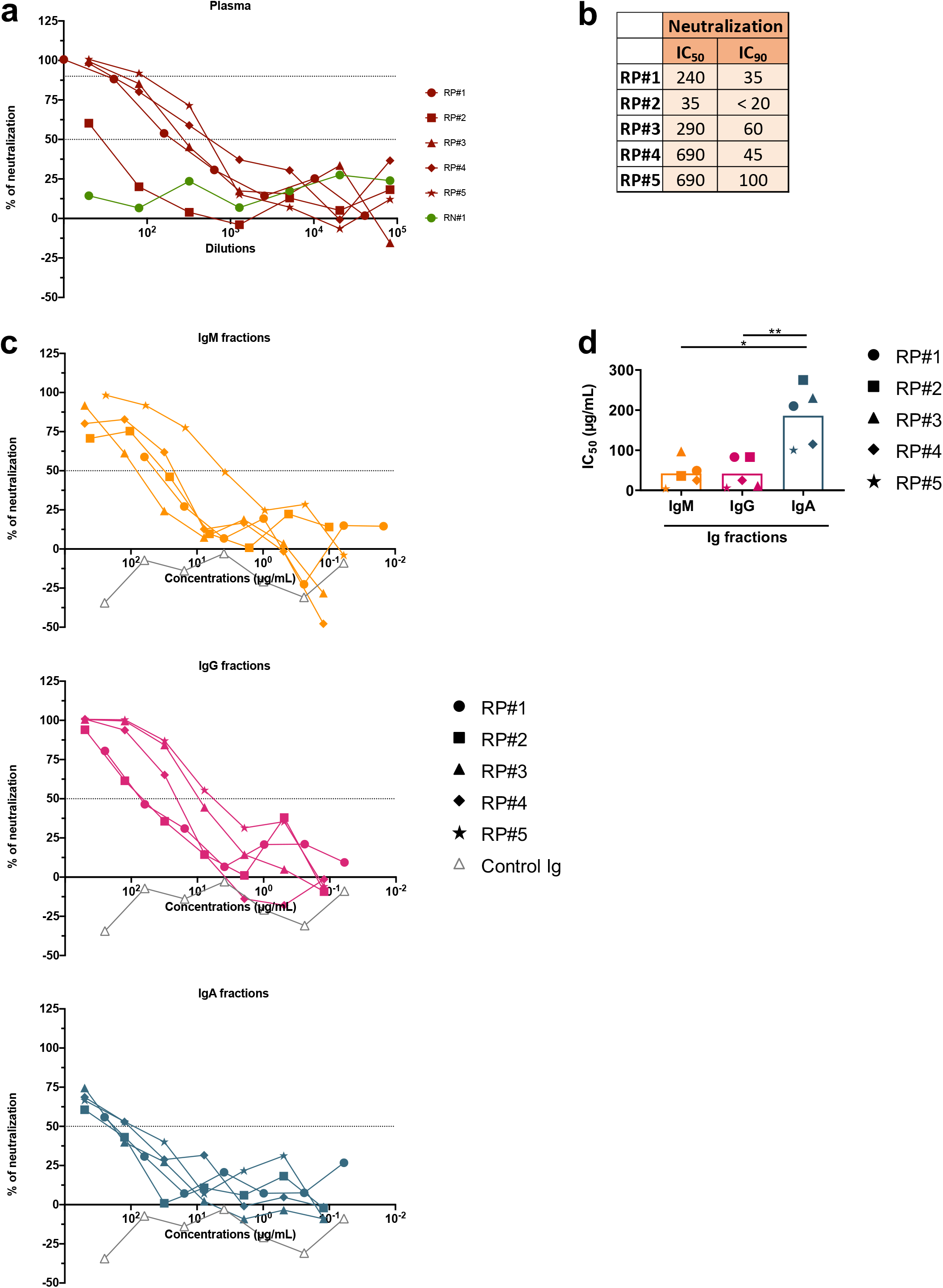
Purified IgM, IgG, and IgA fractions display neutralizing activities against SARS-CoV-2. (a) Neutralization of COV2pp by five COVID-19-infected individual plasma samples (RP#1-5) compared to a specimen from a COVID-19 uninfected individual (RN#1, green filled circles). Plasma samples were tested at 4-fold dilutions from 1:10 to 1:40,960 or 1:20 to 1:81,920. Data are shown as the mean percentage of neutralization. The dotted horizontal lines highlight 50% and 90% neutralization. (b) Reciprocal IC_50_ and IC_90_ neutralization titers of RP#1-5 plasma samples (c) Neutralization of COV2pp by purified IgM, IgG, and IgA fractions from five COVID-19-infected individuals (RP#1-5) compared to a control Ig fraction (gray open triangles). IgA was isolated first from plasma samples by mixing 1:2 diluted plasma with peptide M agarose beads (600 µL/28 mL plasma, InvivoGen #GEL-PDM) for 1.5 hours at room temperature. After washing beads, IgA was eluted with a pH 2.8 buffer (Thermo Scientific #21004) and neutralized with pH 9 Tris buffer. The pass-through plasma sample was collected for IgG enrichment using protein G agarose beads (InvivoGen #GEL-AGG) and subsequently for IgM isolation using a HiTrap IgM column (G.E. Healthcare #17-5110-01). An additional purification step was performed using Protein A Plus mini-spin columns to separate IgG from IgM. The fractions were tested at 4-fold dilutions from 500 to 0.02 µg/mL. Data are shown as the mean percentage of neutralization. The dotted horizontal lines highlight 50% neutralization. (d) IC_50_ of purified IgM, IgG, and IgA fractions from RP#1-5. The statistical significance was determined by a two-tailed Mann-Whitney test (*: p <0.05, **: p <0.01).

## Discussion

Our study demonstrates that IgG1, IgA1 and IgM Abs against SARS-CoV-2 spike and RBD were prevalent in plasma of convalescent COVID-19 patients approximately one to two months after infection. These isotypes were present within 7-8 days after the onset of symptoms. Importantly, all three isotypes showed the capacity to mediate virus neutralization. While regression analyses demonstrated the strongest contributions of IgM and IgG1 to neutralizing activity, direct testing of purified isotype fractions showed that IgA also were able to neutralize, indicating the protective potential of all three major Ig isotypes. These data carry important implications for the use of convalescent plasma and hyperimmunoglobulin as COVID-19 therapeutics, suggesting that their selection would optimally be based on the presence of all of these Ig isotypes.

While all COVID-19 convalescent individuals exhibited neutralization activities reaching >50% and 26 of 28 specimens attained 90% neutralization, neutralization levels were highly variable with IC_50_ and IC_90_ titers ranging over three orders of magnitude. The titers were comparable against the initial Wuhan strain and the currently prevalent D614G strain of SARS-CoV-2. Similarly, the levels of spike- and RBD-binding total Ig and Ig isotypes varied greatly.

A trend toward higher levels of total Ig and each Ig isotype was seen in female compared to male subjects, as reported in another study [36]. Moreover, except for TF#11 (a male elite neutralizer), the median neutralizing IC_90_ titer was higher in females than males, although the difference did not reach significance (data not shown). Sex differences in Ab induction have been observed following influenza vaccination in humans and mice and were shown to result from the impact of sex-related steroids [37]. Whether and to what extent this contributes to the sex differences seen in clinical outcomes of COVID-19 remains to be investigated. Other studies have shown that Ab levels were associated with multiple factors, including time from disease onset [38] and disease severity [14]. However, other than sex, clinical data are not available for the subjects studied here, limiting our analysis only to neutralization and Ig isotypes.

One remarkable finding from our study is that although neutralization titers correlated with binding levels of IgM and IgG1 and not with those of IgA1 or IgA2, purified IgA fractions from convalescent COVID-19 patients exhibited significant neutralizing activities. The importance of this finding is underscored by the data showing that IgA1 was the prominent isotype in some samples such as TF#7 and TF#24 and that IgA1 could be detected early after symptom onset. Data from other studies also support the significance of IgA in that purified IgA fractions exhibited more, or as potent neutralizing activities as purified IgG, and that RBD-binding IgA correlated as strongly as IgG with micro-neutralization titers [39]. IgA were also detected in saliva and bronchoalveolar lavage from COVID-19 patients [40]. Nonetheless, Wang *et al*. reported that plasma IgA monomers were less potent than the plasma IgG and secretory IgA counterparts [41]. In our study, neutralization activities detected in the IgA fractions were mediated mainly by IgA1, the predominant IgA isotype in plasma, and the IC_50_ potency of the IgA fraction was ∼4-fold lower than the potency of IgM and IgG1 fractions. This difference cannot be explained entirely by lower amounts of spike-specific IgA1 in the tested fractions, as estimations using spike-specific monoclonal IgA and IgM Abs yielded similar IgA and IgM concentrations in the respective purified fractions (median of 2 and 2.5 µg/mL respectively). Fine epitope specificities and affinities may differ for IgA, IgM, and IgG to impact neutralization potency, but have yet to be evaluated.

In addition to neutralization, non-neutralizing Ab activities have been implicated in protection from various virus infection through potent Fc-mediated functions such as antibody-dependent cellular cytotoxicity (ADCC), antibody-dependent cellular phagocytosis (ADCP), and complement-mediated lysis; this is reported for HIV, influenza, Marburg, and Ebola viruses [25,42–44]. The Fc activities were not evaluated in our study, and their contribution to protection against SARS-CoV-2 is yet unclear [45,46]. A recent study demonstrated enrichment of spike-specific IgM and IgA1 Abs and spike-specific phagocytic and antibody-dependent complement deposition (ADCD) activity in plasma of individuals who recovered from SARS-CoV-2 infection, while nucleocapsid-specific IgM and IgA2 responses and nucleocapsid-specific ADCD activity were features enriched in deceased patients [47]. DNA vaccines expressing full-length and truncated spike proteins could curtail SARS-CoV-2 infection in the respiratory tract by varying degrees in rhesus macaques. This virus reduction correlated with levels of neutralization and also with Fc-mediated effector functions such as ADCD [45]. Interestingly, these DNA vaccines elicited spike- and RBD-specific IgG1, IgG2, IgG3, IgA, and IgM Abs, and similar to our findings, neutralization correlated most strongly with IgM. Adenovirus serotype 26 vaccine vectors encoding seven SARS-CoV-2 spike variants also showed varying protection levels, and virus reduction correlated best with neutralizing titers together with IgM binding levels, FcγRII-binding, and ADCD responses [48]. Defining the full functional potential of Abs against SARS-CoV-2—including neutralizing, non-neutralizing, and enhancing activities—are vital for determining the optimal Ab treatment modalities against COVID-19 and the potential efficacy of COVID-19 vaccine candidates.

When we examined plasma specimens collected within 7-8 days after COVID-19 symptom onset, we detected IgG and IgA against spike and RBD, as well as IgM. This is consistent with published reports showing that 100% of COVID-19-infected individuals developed IgG within 19 days after symptom onset and that IgG and IgM seroconversion could occur simultaneously [14]. IgA were also found early after infection (4-6 days after symptom onset) and increased over time [13,18,40]. These studies suggest that measuring total Ig, rather than IgG, could contribute to improved outcomes for early disease diagnosis. We found no correlation between the levels of different isotypes in the specimens examined in our study (data not shown). Of note, IgA presence early during acute infection may suggest the potential contribution of natural IgA, which, similar to natural IgM, arises spontaneously from innate B1 cells to provide the initial humoral responses before the induction of adaptive classical B cells [49].

In summary, this study demonstrates that spike- and RBD-specific IgM, IgG1, and IgA1 are produced by all or almost all analyzed COVID-19 convalescent subjects and can be detected at early stages of infection. The plasma samples of convalescent individuals also display neutralization activities mediated by IgM, IgG, and IgA1, although neutralization titers correlated more strongly with IgM and IgG levels. The contribution of IgM, IgG, and IgA to SARS-CoV-2-neutralizing activities demonstrates their importance in the efficacy of passively transferred Abs for SARS-CoV-2 treatment.

## Data Availability

The raw data that support the findings of this study are available from the corresponding author upon request.

## Acknowledgments

We thank Dr. Florian Krammer, Dr. Viviana Simon, and Dr. Rebecca Powell for donation of samples and reagents, and all the donors for their contribution to the research.

## Author contributions

J.K., S.W., G.E-A., S.Z-P., and C.E.H. wrote and edited the manuscript. S.W., J.K., C.E.H., and S.Z-P. designed the experiments. J.K., S.W., V.I., X.L. performed the experiments and collected the data. J.K., A.N., S.Z-P. and C.E.H. analyzed the data. K.Y.O., C.S., S.I., C-T.H., F.A., and B.L. provided protocols, antigens, cells and pseudovirus stocks. G.E-A., I.B., S.A., J.C.B., E.M.K., J.S., S.L., D.J., and M.B-G. provided specimens. All authors read and approved the final manuscript.

## Figure Legends

**Supplementary Fig. 1**. Spearman correlations of (a) spike-specific or (b) RBD-specific total Ig MFI values from two independent experiments to show the degree of assay reproducibility.

**Supplementary Fig. 2**. Spearman correlations of the area under the curves (AUCs) of (a) spike- or (b) RBD-specific total Ig versus total Ig MFI values at a 1:200 dilution.

**Supplementary Fig. 3**. Isotyping validation was performed by coating Luminex beads with IgG1, IgG2, IgG3, IgG4, IgA1, IgA2, and IgM myeloma proteins and detecting each with eight different secondary Abs against total Ig, IgM, IgG1, IgG2, IgG3, IgG4, IgA1 and IgA2. The data are shown as mean MFI + SD of duplicate.

**Supplementary Fig. 4**. Spearman correlations between spike-specific versus RBD-specific total Ig, IgM, IgG1, IgG2, IgG3, IgG4, IgA1, or IgA2 MFI values.

**Supplementary Fig. 5**. Violin plots of (a) spike-specific or (b) RBD-specific total Ig, IgM, IgG1, and IgA1 levels from nine COVID-19 convalescent female (F) and 15 male (M) subjects. The statistical significance was determined by a two-tailed Mann-Whitney test (ns: non-significant: p > 0.05).

**Supplementary Fig. 6. Induction of IgA1 and IgG1 along with IgM early after disease onset**. Kinetics of induction of spike-specific (left panel) or RBD-specific (right panel) total Ig, IgM, IgG1, IgG2, IgG3, IgG4, IgA1, and IgA2 from two COVID-19 patients. Longitudinal samples from each patient were tested at a dilution of 1:200 in parallel with all negative samples and data are shown as mean MFI + SD of duplicate measurements from at least two experiments. The dotted red line represents the cut-off value calculated as the mean of 12 pre-pandemic samples + 3 SD from Fig. 1.

**Supplementary Fig. 7. Enrichment of spike-specific (a) IgM, (b) IgG, and (c) IgA in purified fractions from RP#1-5 and RN#1**. Each purified isotype fraction from plasma was measured for the presence of IgM, IgG1, IgG2, IgG3, IgG4, IgA1, and IgA2 Abs using the isotyping method validated in Supplementary Fig. 3.

