## Supplemental Figures 1-7 for "Role of IgM and IgA Antibodies in the Neutralization of SARS-CoV-2"

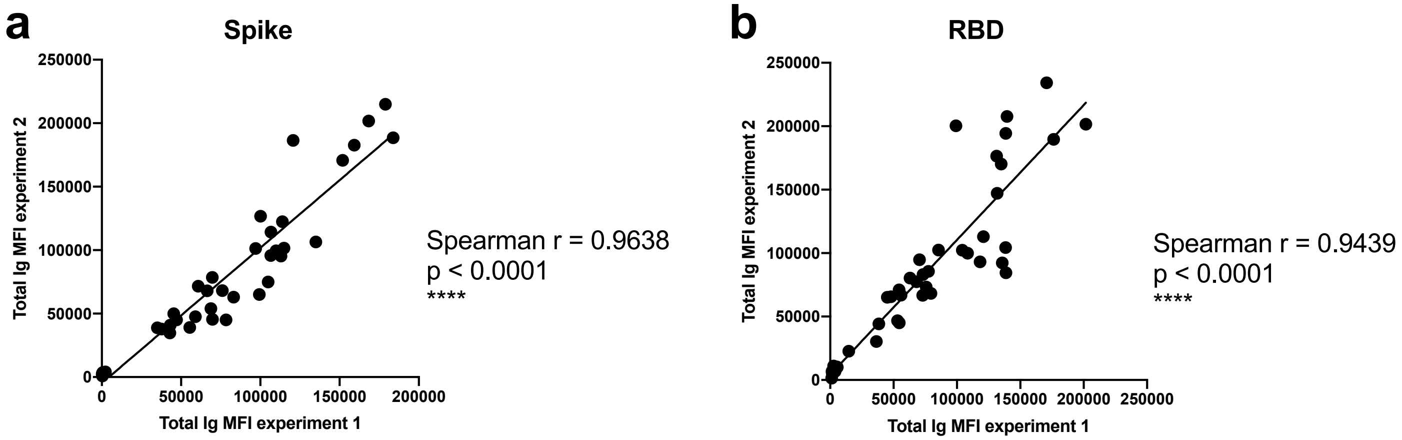
a
Spike
Total Ig MFI experiment 2
Total Ig MFI experiment 1
0
50000
100000
150000
200000
250000
Spearman r = 0.9638
p < 0.0001
****
b
RBD
Total Ig MFI experiment 2
Total Ig MFI experiment 1
0
50000
100000
150000
200000
250000
Spearman r = 0.9439
p < 0.0001
****

**Supplementary Fig. 1.** Spearman correlations of (a) spike-specific or (b) RBD-specific total Ig MFI values from two independent experiments to show the degree of assay reproducibility.

a

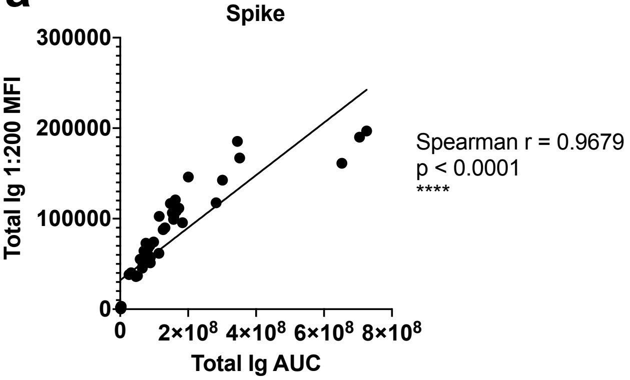
Spike
Total Ig 1:200 MFI
300000
200000
100000
0
0
2×10^8
4×10^8
6×10^8
8×10^8
Total Ig AUC
Spearman r = 0.9679
p < 0.0001
****

b

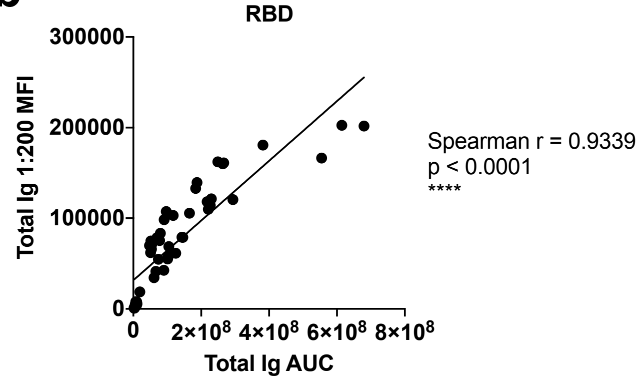
RBD
Total Ig 1:200 MFI
300000
200000
100000
0
0
2×10^8
4×10^8
6×10^8
8×10^8
Total Ig AUC
Spearman r = 0.9339
p < 0.0001
****

**Supplementary Fig. 2.** Spearman correlations of the area under the curves (AUCs) of (a) spike- or (b) RBD-specific total Ig versus total Ig MFI values at a 1:200 dilution.

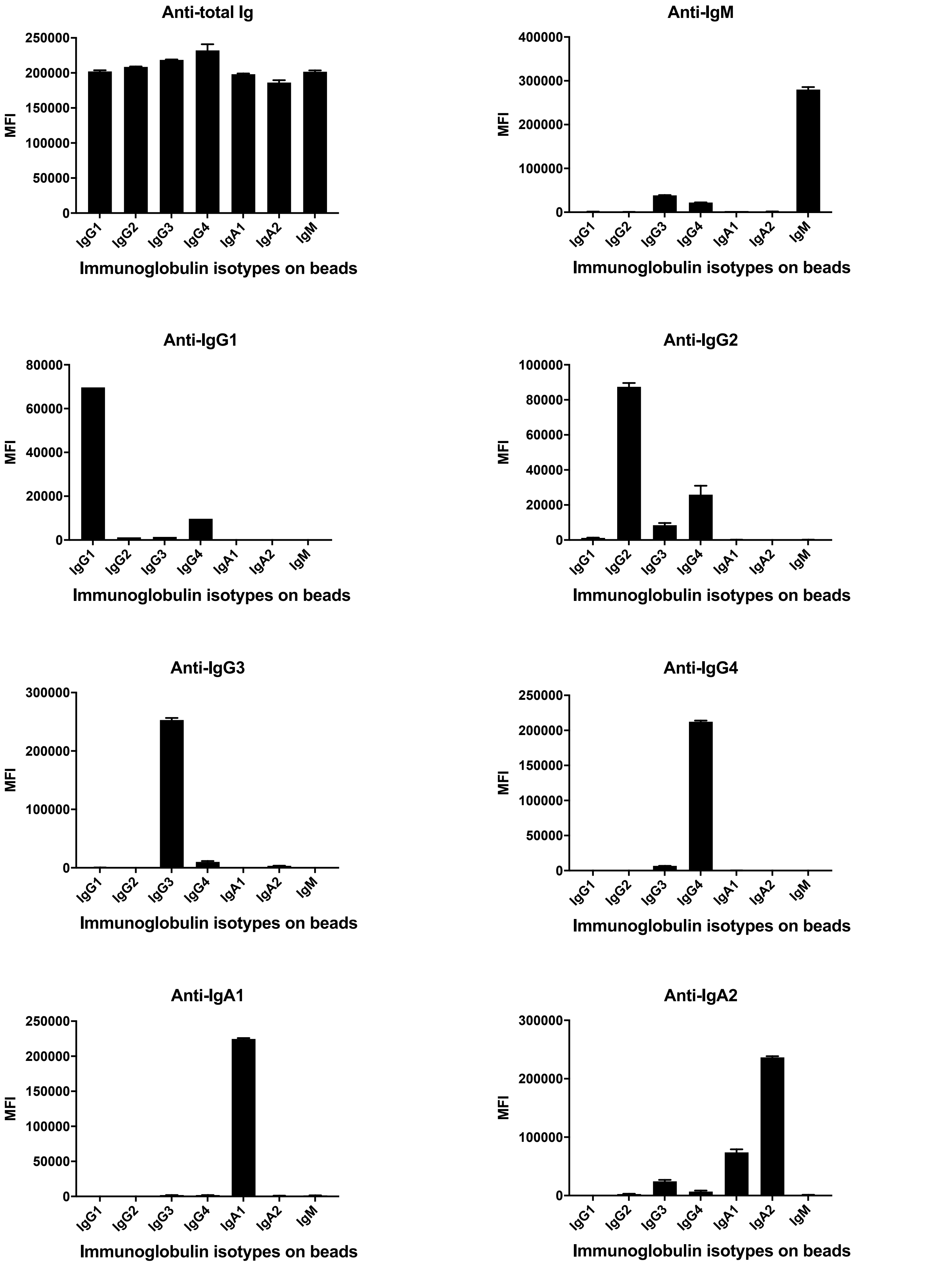

Anti-total Ig
250000
200000
150000
100000
50000
0
MFI
IgG1
IgG2
IgG3
IgG4
IgA1
IgA2
IgM
Immunoglobulin isotypes on beads
Anti-IgM
400000
300000
200000
100000
0
MFI
IgG1
IgG2
IgG3
IgG4
IgA1
IgA2
IgM
Immunoglobulin isotypes on beads
Anti-IgG1
80000
60000
40000
20000
0
MFI
IgG1
IgG2
IgG3
IgG4
IgA1
IgA2
IgM
Immunoglobulin isotypes on beads
Anti-IgG2
100000
80000
60000
40000
20000
0
MFI
IgG1
IgG2
IgG3
IgG4
IgA1
IgA2
IgM
Immunoglobulin isotypes on beads
Anti-IgG3
300000
200000
100000
0
MFI
IgG1
IgG2
IgG3
IgG4
IgA1
IgA2
IgM
Immunoglobulin isotypes on beads
Anti-IgG4
250000
200000
150000
100000
50000
0
MFI
IgG1
IgG2
IgG3
IgG4
IgA1
IgA2
IgM
Immunoglobulin isotypes on beads
Anti-IgA1
250000
200000
150000
100000
50000
0
MFI
IgG1
IgG2
IgG3
IgG4
IgA1
IgA2
IgM
Immunoglobulin isotypes on beads
Anti-IgA2
300000
200000
100000
0
MFI
IgG1
IgG2
IgG3
IgG4
IgA1
IgA2
IgM
Immunoglobulin isotypes on beads

**Supplementary Fig. 3.** Isotyping validation was performed by coating Luminex beads with IgG1, IgG2, IgG3, IgG4, IgA1, IgA2, and IgM myeloma proteins and detecting each with eight different secondary Abs against total Ig, IgM, IgG1, IgG2, IgG3, IgG4, IgA1 and IgA2. The data are shown as mean MFI + SD of duplicate.

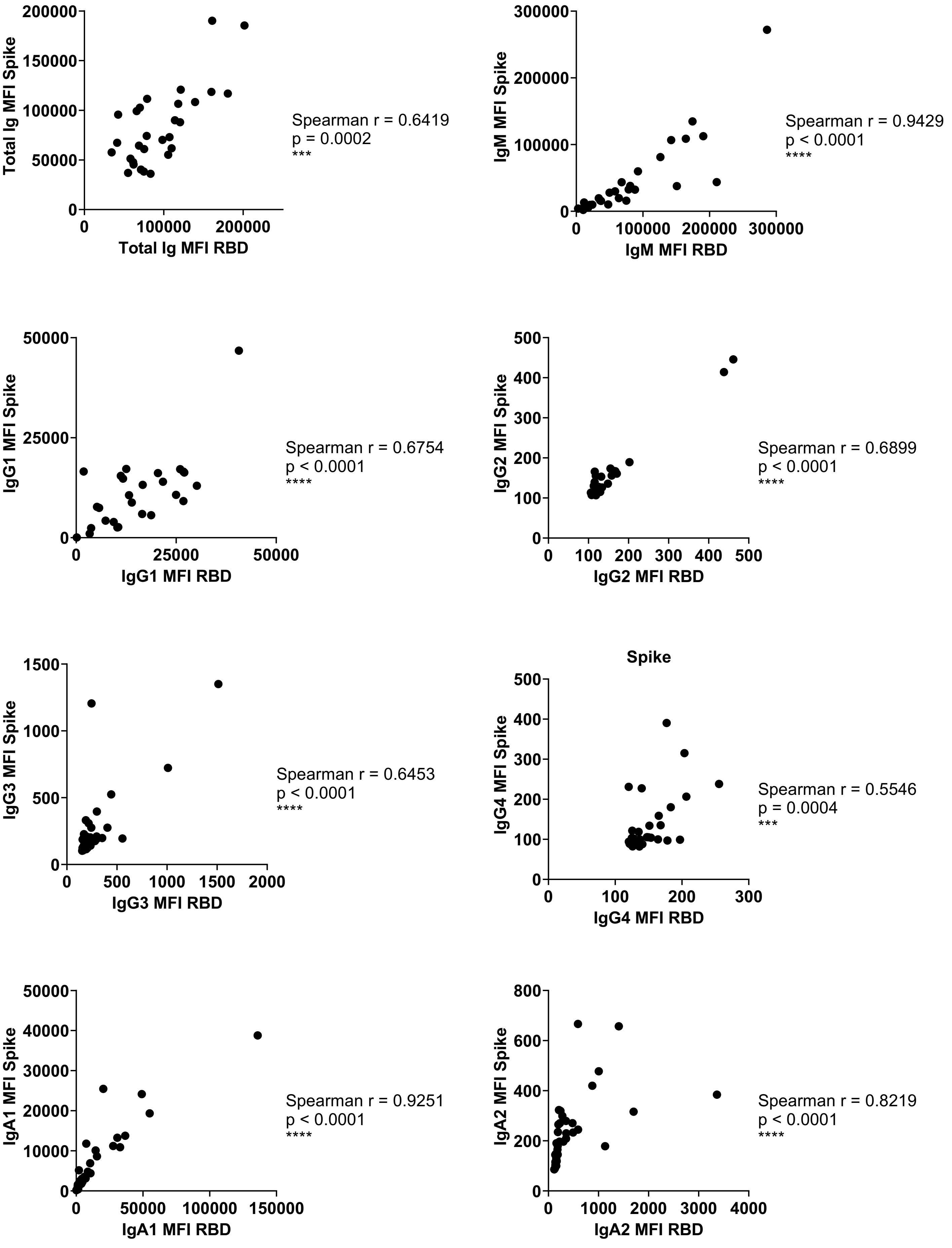
Total Ig MFI Spike
Total Ig MFI RBD
Spearman r = 0.6419
p = 0.0002
***
IgM MFI Spike
IgM MFI RBD
Spearman r = 0.9429
p < 0.0001
****
IgG1 MFI Spike
IgG1 MFI RBD
Spearman r = 0.6754
p < 0.0001
****
IgG2 MFI Spike
IgG2 MFI RBD
Spearman r = 0.6899
p < 0.0001
****
IgG3 MFI Spike
IgG3 MFI RBD
Spearman r = 0.6453
p < 0.0001
****
Spike
IgG4 MFI Spike
IgG4 MFI RBD
Spearman r = 0.5546
p = 0.0004
***
IgA1 MFI Spike
IgA1 MFI RBD
Spearman r = 0.9251
p < 0.0001
****
IgA2 MFI Spike
IgA2 MFI RBD
Spearman r = 0.8219
p < 0.0001
****

**Supplementary Fig. 4.** Spearman correlations between spike-specific versus RBD-specific total Ig, IgM, IgG1, IgG2, IgG3, IgG4, IgA1, or IgA2 MFI values.

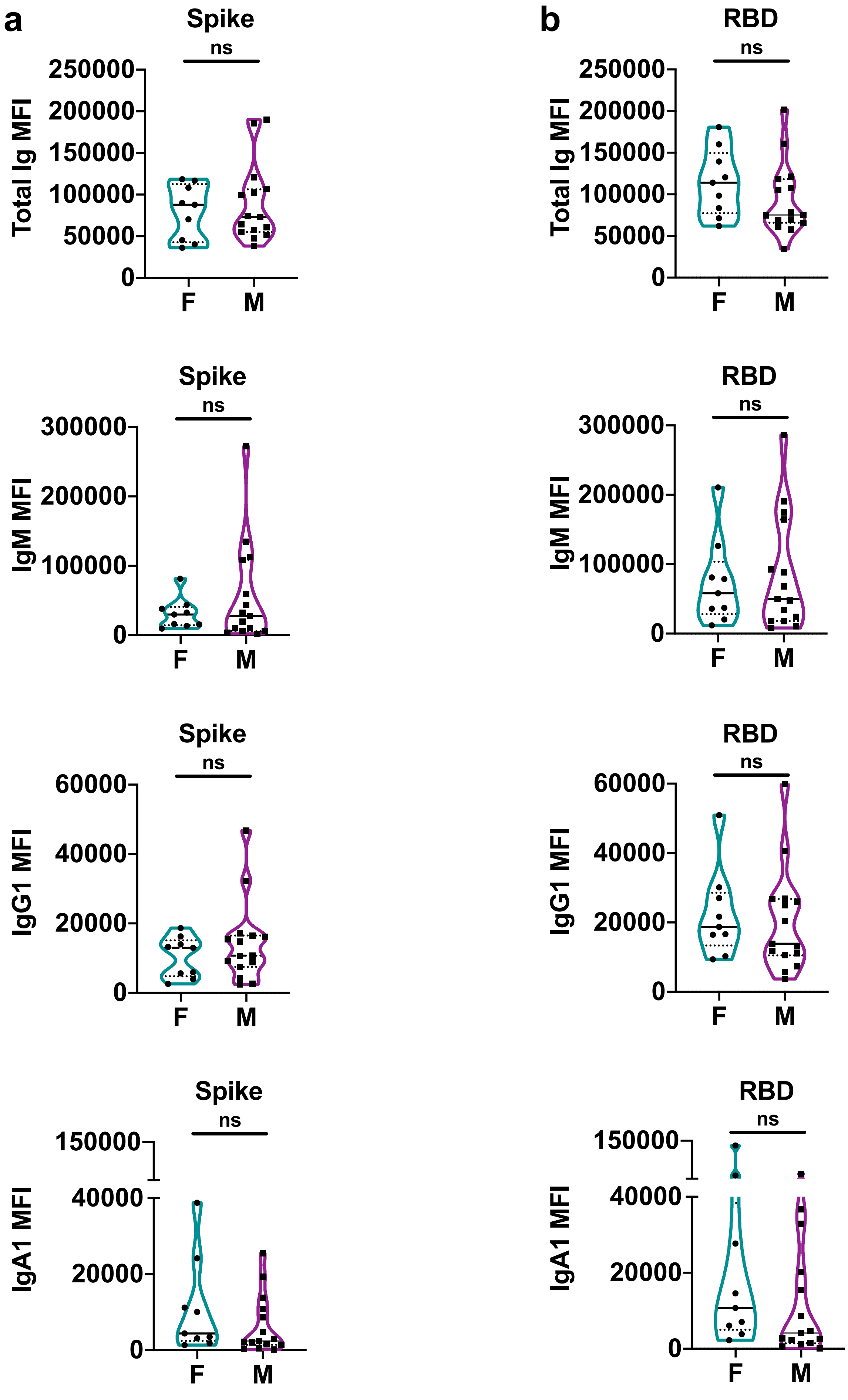
a
b
Spike
RBD
ns
Total Ig MFI
IgM MFI
IgG1 MFI
IgA1 MFI
250000
200000
150000
100000
50000
300000
60000
40000
20000
0
F
M

**Supplementary Fig. 5.** Violin plots of (a) spike-specific or (b) RBD-specific total Ig, IgM, IgG1, and IgA1 levels from nine COVID-19 convalescent female (F) and 15 male (M) subjects. The statistical significance was determined by a two-tailed Mann-Whitney test (ns: non-significant: $p > 0.05$).

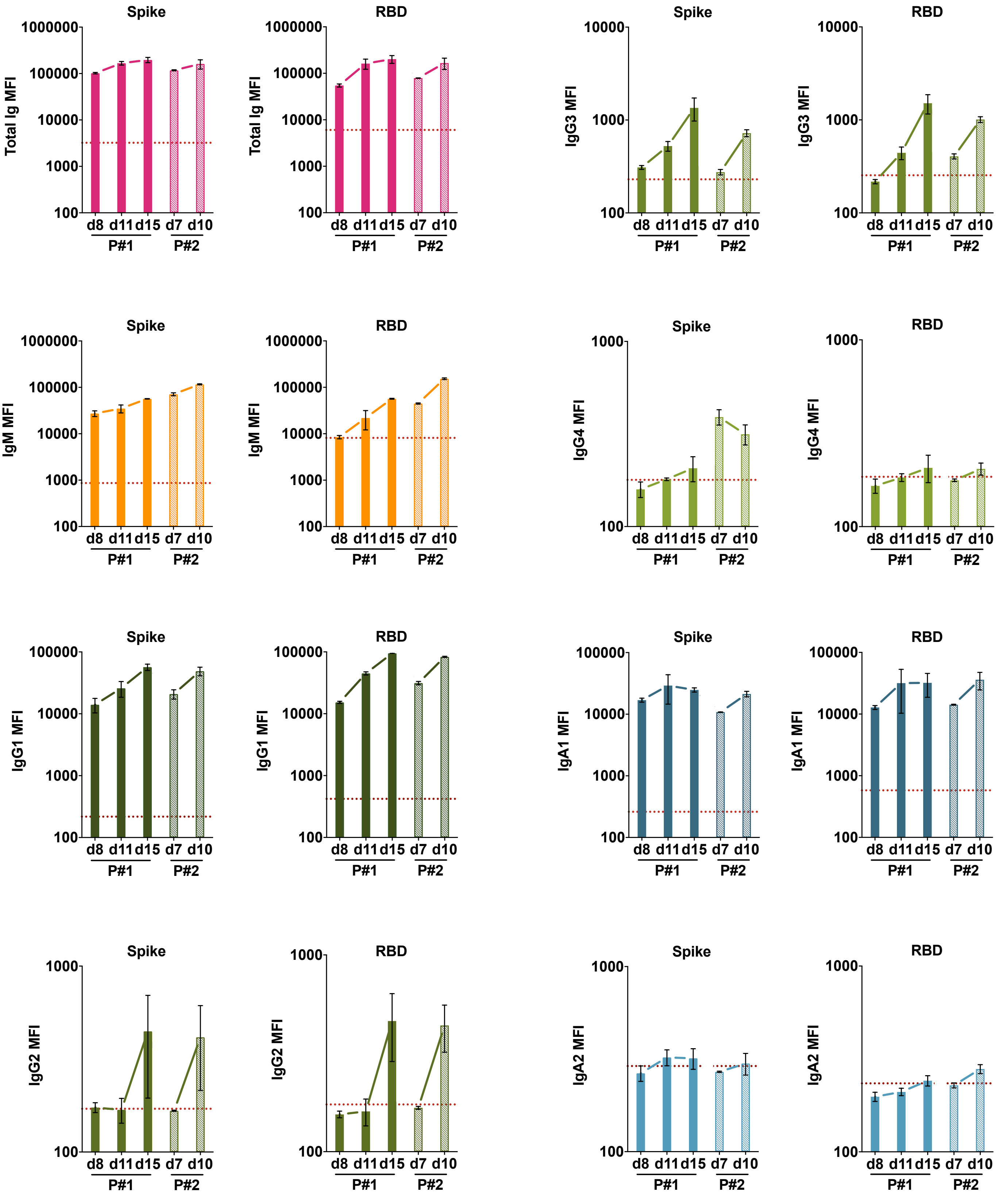
Spike
Total Ig MFI
1000000
100000
10000
1000
100
d8 d11 d15 d7 d10
P#1 P#2
RBD
Total Ig MFI
1000000
100000
10000
1000
100
d8 d11 d15 d7 d10
P#1 P#2
Spike
IgG3 MFI
10000
1000
100
d8 d11 d15 d7 d10
P#1 P#2
RBD
IgG3 MFI
10000
1000
100
d8 d11 d15 d7 d10
P#1 P#2
Spike
IgM MFI
1000000
100000
10000
1000
100
d8 d11 d15 d7 d10
P#1 P#2
RBD
IgM MFI
1000000
100000
10000
1000
100
d8 d11 d15 d7 d10
P#1 P#2
Spike
IgG4 MFI
1000
100
d8 d11 d15 d7 d10
P#1 P#2
RBD
IgG4 MFI
1000
100
d8 d11 d15 d7 d10
P#1 P#2
Spike
IgG1 MFI
100000
10000
1000
100
d8 d11 d15 d7 d10
P#1 P#2
RBD
IgG1 MFI
100000
10000
1000
100
d8 d11 d15 d7 d10
P#1 P#2
Spike
IgA1 MFI
100000
10000
1000
100
d8 d11 d15 d7 d10
P#1 P#2
RBD
IgA1 MFI
100000
10000
1000
100
d8 d11 d15 d7 d10
P#1 P#2
Spike
IgG2 MFI
1000
100
d8 d11 d15 d7 d10
P#1 P#2
RBD
IgG2 MFI
1000
100
d8 d11 d15 d7 d10
P#1 P#2
Spike
IgA2 MFI
1000
100
d8 d11 d15 d7 d10
P#1 P#2
RBD
IgA2 MFI
1000
100
d8 d11 d15 d7 d10
P#1 P#2

**Supplementary Fig. 6. Induction of IgA1 and IgG1 along with IgM early after disease onset.** Kinetics of induction of spike-specific (left panel) or RBD-specific (right panel) total Ig, IgM, IgG1, IgG2, IgG3, IgG4, IgA1, and IgA2 from two COVID-19 patients. Longitudinal samples from each patient were tested at a dilution of 1:200 in parallel with all negative samples and data are shown as mean MFI + SD of duplicate measurements from at least two experiments. The dotted red line represents the cut-off value calculated as the mean of 12 pre-pandemic samples + 3 SD from Fig. 1.

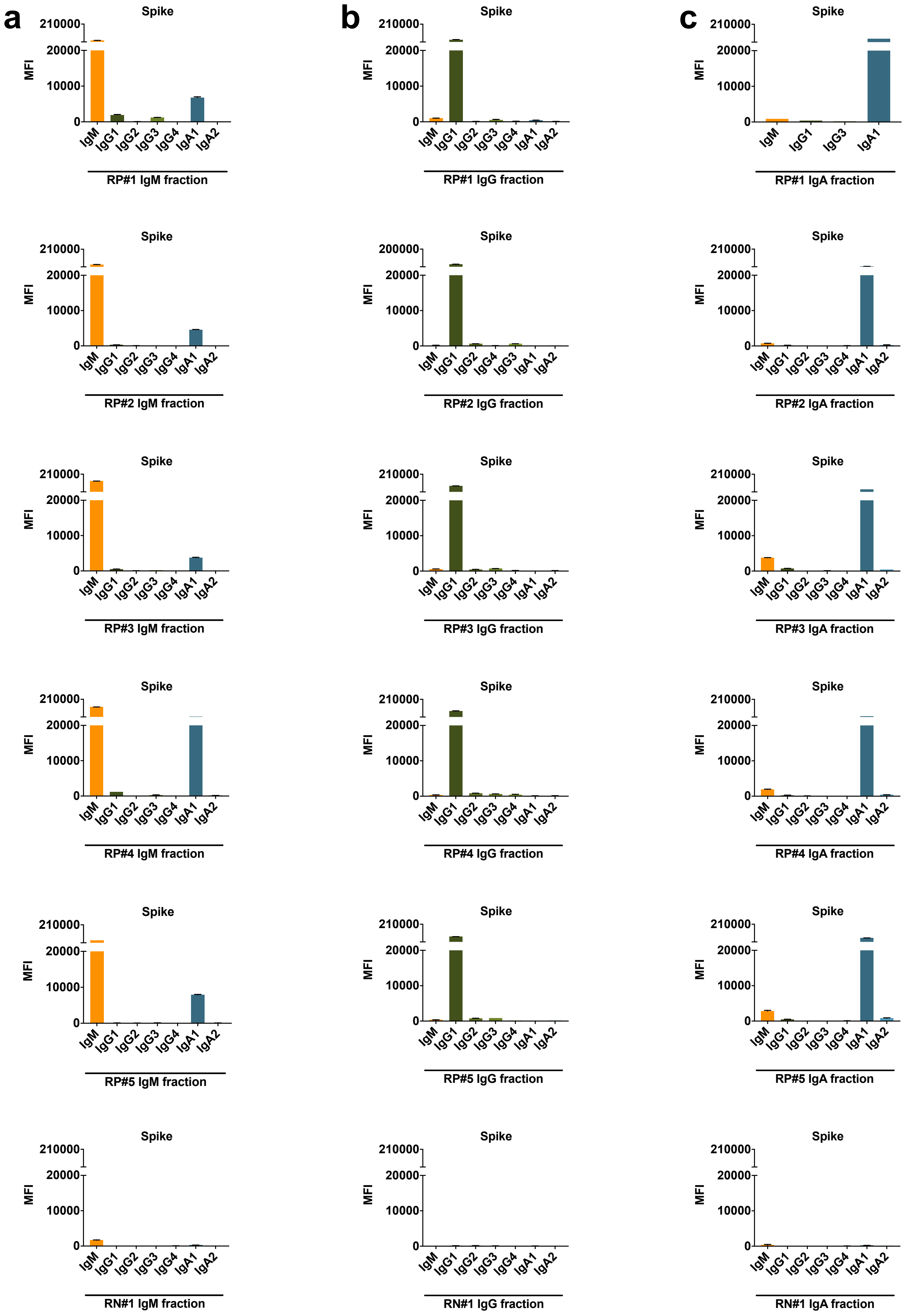
a
b
c
Spike
MFI
210000
20000
10000
0
IgM IgG1 IgG2 IgG3 IgG4 IgA1 IgA2
RP#1 IgM fraction
RP#1 IgG fraction
IgM IgG1 IgG3 IgA1
RP#1 IgA fraction
RP#2 IgM fraction
200000
IgM IgG1 IgG2 IgG4 IgG3 IgA1 IgA2
RP#2 IgG fraction
RP#2 IgA fraction
RP#3 IgM fraction
RP#3 IgG fraction
RP#3 IgA fraction
RP#4 IgM fraction
RP#4 IgG fraction
RP#4 IgA fraction
RP#5 IgM fraction
RP#5 IgG fraction
RP#5 IgA fraction
RN#1 IgM fraction
RN#1 IgG fraction
RN#1 IgA fraction

**Supplementary Fig. 7. Enrichment of spike-specific (a) IgM, (b) IgG, and (c) IgA in purified fractions from RP#1-5 and RN#1.** Each purified isotype fraction from plasma was measured for the presence of IgM, IgG1, IgG2, IgG3, IgG4, IgA1, and IgA2 Abs using the isotyping method validated in Supplementary Fig. 3.
