## Supplemental Table 1 for "Role of IgM and IgA Antibodies in the Neutralization of SARS-CoV-2"

**Supplementary Table 1. Summary of COVID-19-positive and -negative samples examined in the study.**

| | Specimen identifier | Biospecimen | Age range | Sex | Transfused to patient? | Date of plasma donation | Days between sera testing and plasma donation | SARS-CoV-2 PCR result | Symptoms suggestive of COVID-19 | Days post symptom onset and serum collection | COVID-19 severity |
|---|---|---|---|---|---|---|---|---|---|---|---|
| Individuals with acute infection | **P#1a** | serum | unknown | unknown | no | n/a | n/a | positive | yes | 8 | severe |
| | **P#1b** | serum | unknown | unknown | no | n/a | n/a | positive | yes | 11 | severe |
| | **P#1c** | serum | unknown | unknown | no | n/a | n/a | positive | yes | 15 | severe |
| | **P#2a** | serum | unknown | unknown | no | n/a | n/a | positive | yes | 7 | severe |
| | **P#2b** | serum | unknown | unknown | no | n/a | n/a | positive | yes | 10 | severe |
| | **P#3** | serum | unknown | unknown | no | n/a | n/a | positive | yes | 22 | mild |
| | **P#4** | serum | unknown | unknown | no | n/a | n/a | inconclusive | yes | 21 | mild |
| COVID-19-convalescent individuals | **P#5** | serum | unknown | unknown | no | n/a | n/a | positive | unknown | unknown | unknown |
| | **P#6** | serum | unknown | unknown | no | n/a | n/a | positive | unknown | unknown | unknown |
| | **P#7** | serum | unknown | unknown | no | n/a | n/a | positive | unknown | unknown | unknown |
| | **P#8** | serum | unknown | unknown | no | n/a | n/a | positive | unknown | unknown | unknown |
| | **TF#1** | plasma | 50-55 | F | yes | 26/03/2020 - 07/04/2020 | 2 | unknown | unknown | unknown | unknown |
| | **TF#2** | plasma | 30-35 | M | yes | | 4 | unknown | unknown | unknown | unknown |
| | **TF#3** | plasma | 25-30 | M | yes | | 4 | unknown | unknown | unknown | unknown |
| | **TF#4** | plasma | 30-35 | F | yes | | 3 | unknown | unknown | unknown | unknown |
| | **TF#5** | plasma | 45-50 | M | yes | | 6 | unknown | unknown | unknown | unknown |
| | **TF#6** | plasma | 15-20 | F | yes | | 3 | unknown | unknown | unknown | unknown |
| | **TF#7** | plasma | 35-40 | F | yes | | 2 | unknown | unknown | unknown | unknown |
| | **TF#8** | plasma | 50-55 | M | yes | | 1 | unknown | unknown | unknown | unknown |
| | **TF#9** | plasma | 35-40 | F | yes | | 5 | unknown | unknown | unknown | unknown |
| | **TF#10** | plasma | 35-40 | M | yes | | 7 | unknown | unknown | unknown | unknown |
| | **TF#11** | plasma | 50-55 | M | yes | | 6 | unknown | unknown | unknown | unknown |
| | **TF#12** | plasma | 25-30 | F | yes | | 4 | unknown | unknown | unknown | unknown |
| | **TF#13** | plasma | 30-35 | M | yes | | 3 | unknown | unknown | unknown | unknown |
| | **TF#14** | plasma | 25-30 | M | yes | | 3 | unknown | unknown | unknown | unknown |
| | **TF#15** | plasma | 35-40 | M | yes | | 3 | unknown | unknown | unknown | unknown |
| | **TF#16** | plasma | 45-50 | M | yes | | 3 | unknown | unknown | unknown | unknown |
| | **TF#17** | plasma | 25-30 | M | yes | | 4 | unknown | unknown | unknown | unknown |
| | **TF#18** | plasma | 30-35 | F | yes | | 5 | unknown | unknown | unknown | unknown |
| | **TF#19** | plasma | 30-35 | M | yes | | 4 | unknown | unknown | unknown | unknown |
| | **TF#20** | plasma | 25-30 | F | yes | | 3 | unknown | unknown | unknown | unknown |
| | **TF#21** | plasma | 25-30 | M | yes | | 5 | unknown | unknown | unknown | unknown |
| | **TF#22** | plasma | 30-35 | M | yes | | 6 | unknown | unknown | unknown | unknown |
| | **TF#23** | plasma | 30-35 | unknown | yes | | 6 | unknown | unknown | unknown | unknown |
| | **TF#24** | plasma | 30-35 | M | yes | | 3 | unknown | unknown | unknown | unknown |
| | **TF#25** | plasma | 50-55 | F | yes | | 6 | unknown | unknown | unknown | unknown |
| Pre-pandemic controls | **N#1** | serum | unknown | unknown | n/a | n/a | n/a | n/a | n/a | n/a | n/a |
| | **N#2** | serum | unknown | unknown | n/a | n/a | n/a | n/a | n/a | n/a | n/a |
| | **N#3** | serum | unknown | unknown | n/a | n/a | n/a | n/a | n/a | n/a | n/a |
| Contemporaneous COVID-19-negative | **N#4** | plasma | unknown | unknown | n/a | n/a | n/a | n/a | n/a | n/a | n/a |
| | **N#5** | plasma | unknown | unknown | n/a | n/a | n/a | n/a | n/a | n/a | n/a |
| | **N#6** | plasma | unknown | unknown | n/a | n/a | n/a | n/a | n/a | n/a | n/a |
| | **N#7** | plasma | unknown | unknown | n/a | n/a | n/a | n/a | n/a | n/a | n/a |
| | **N#8** | plasma | unknown | unknown | n/a | n/a | n/a | n/a | n/a | n/a | n/a |
| | **N#9** | plasma | unknown | unknown | n/a | n/a | n/a | n/a | n/a | n/a | n/a |
| | **N#10** | plasma | unknown | unknown | n/a | n/a | n/a | n/a | n/a | n/a | n/a |
| | **N#11** | plasma | unknown | unknown | n/a | n/a | n/a | n/a | n/a | n/a | n/a |
| | **N#12** | plasma | unknown | unknown | n/a | n/a | n/a | n/a | n/a | n/a | n/a |
| | **N#13** | plasma | unknown | unknown | n/a | n/a | n/a | n/a | n/a | n/a | n/a |
| Pre-pandemic controls | **N#14** | plasma | 60-65 | M | n/a | n/a | n/a | n/a | n/a | n/a | n/a |
| | **N#15** | plasma | 65-70 | M | n/a | n/a | n/a | n/a | n/a | n/a | n/a |
| | **N#16** | plasma | 50-55 | M | n/a | n/a | n/a | n/a | n/a | n/a | n/a |
| | **N#17** | plasma | 65-70 | M | n/a | n/a | n/a | n/a | n/a | n/a | n/a |
| | **N#18** | plasma | 65-70 | M | n/a | n/a | n/a | n/a | n/a | n/a | n/a |
| | **N#19** | plasma | 60-65 | M | n/a | n/a | n/a | n/a | n/a | n/a | n/a |
| | **N#20** | plasma | 65-70 | M | n/a | n/a | n/a | n/a | n/a | n/a | n/a |
| | **N#21** | plasma | 50-55 | M | n/a | n/a | n/a | n/a | n/a | n/a | n/a |
| | **N#22** | plasma | 55-60 | M | n/a | n/a | n/a | n/a | n/a | n/a | n/a |

F: female, M: male, n/a: not applicable.
